# Clinical characteristics and serotype association of dengue and dengue like illness in Pakistan

**DOI:** 10.1101/2024.10.02.24314748

**Authors:** Najeeha Talat Iqbal, Kumail Ahmed, Aqsa Khalid, Kehkashan Imtiaz, Qamreen Mumtaz Ali, Tania Munir, Syed Faisal Mahmood, Unab Khan, Badar Afzal, Farah Qamar, Jesse J. Waggoner, Hannah Fenelon, Helene McOwen, Erum Khan, Peter Rabinowitz, Wesley C. Van Voorhis

**Author notes:** Corresponding Author: **Najeeha Talat Iqbal**, (NTA).

## Abstract

**Background:** Pakistan has been an endemic country for dengue virus since 1994, with a significant increase in cases reported in 2022 largely due to heavy rainfall and flooding. All four serotypes of the dengue virus (DENV) are present in Pakistan, with DENV 1 and DENV 2 being the most prevalent. The current study aimed to explore the clinical presentations and features of dengue fever in a tertiary care hospital.

**Methodology:** We enrolled and studied 349 cases of suspected and confirmed dengue presenting for care at the Aga Khan University Hospital in Karachi between June 2021 and November 2023. Collected data on cases including clinical symptoms and laboratory results including qRT-PCR and serotype characterization.

**Findings:** The majority of subjects enrolled (75%) had mild disease without warning signs, while 11% exhibited warning signs, 1.4% had severe dengue, and 12.6 % had no dengue diagnosis. Patients with severe dengue (SD) had significantly higher levels of liver enzymes (AST and ALT) compared to those with non-severe dengue (NSD) (AST; p=0.024 and ALT; p=0.047). Additionally, a higher grade of thrombocytopenia was significantly associated with hospitalization (p=0.0008), and prolonged illness (p=0.03). Both Platelet (p<0.0001) and WBC counts (p=0.001) were significantly lower in dengue PCR-positive patients in comparison to Dengue PCR-negative. Among those tested for dengue serotypes, DENV 1 (34%) and DENV 2 (45%) emerged as the predominant serotypes, with mixed infections accounting for 17%. The sensitivity of clinical diagnosis was found to be 87.25% and specificity of 68.35%. qRT-PCR detected 43.5% of cases with viral fever initially screened negative.

**Conclusion:** Epidemiology of dengue fever during a widespread outbreak in 2022 showed a predominance of DENV 1 and DENV 2 serotypes with milder phenotype of viral illness. Screening with rapid tests requires further confirmation by molecular assay in cases with dengue and dengue-like illness.

## Introduction

Dengue is prevalent in tropical and subtropical regions and is transmitted by Aedes aegypti mosquitoes(6). While most cases are self-limited, some can progress to the life-threatening hemorrhagic shock syndrome. Dengue is caused by four distinct serotypes of the dengue virus such as DENV 1, DENV 2, DENV 3, and DENV 4 (7). These four serotypes have been reported in different outbreaks of dengue infection in Pakistan with DENV 2 and DENV 3 to be the most prevalent serotypes (7). A single serotype infection provides relative protection against that specific serotype, whereas reinfection with other serotypes may lead to more severe illness(8). The distribution of dengue serotypes in Pakistan is uneven, with DENV 2 being reported in 1994 and subsequently in 2005 and 2006, along with DENV 3(8). DENV 1 and DENV 2 were identified as the most prevalent serotypes during the 2022 outbreak in Pakistan(9). The studies conducted in neighboring countries such as India, Indonesia, and Malaysia also reported the severity of the dengue infection association with concurrent infection of DENV 1/ DENV 2 and DENV 2/ DENV 3 (10) (11).

In 2022, torrential monsoon rains accompanied by flooding affected many villages and households in Pakistan, with rain fall 175% higher than the annual average over the past 60 years, and as high as 726% of the annual average in Sindh province. This severe event caused damage to crops, livestock, and infrastructure and had a huge toll on the communities with low socio-economic status. These flood conditions were accompanied by an upsurge in waterborne and vector borne diseases, including a rapid spread of dengue infection(3) (4). The World Health Organization (WHO) report on the 2022 dengue outbreak in Pakistan, concluded that there was a total of 25,932 confirmed cases and 62 fatalities among individuals seeking medical assistance for health concerns (5).

The Aga Khan University Hospital in Karachi, Pakistan is a tertiary care center that receives cases of dengue from the Karachi metropolitan area; a highly urbanized city with suboptimal sanitation and weak infrastructure of society, open garbage, and leaky pipelines provides an ideal breeding environment for vectors. The fact that all four serotypes were circulating during the outbreak of 2022 provided an opportunity to compare clinical aspects of the serotypes receiving care in the same facility.

It is also important to acknowledge the possibility of other circulating viruses like Zika virus and Chikungunya viruses which share a common host (human) and mosquito vectors (primarily Aedes aegypti) and have similar biological and ecological factors leading to epidemiological synergy (12, 13).

We therefore surveyed cases of confirmed and suspected dengue that were hospitalized or treated as out-patients between 2021-2023, which captured the period of the 2022 outbreak, allowing us to highlight the clinical features and circulation of the different dengue serotypes, as well as the sensitivity of PCR compared to clinical diagnosis.

## 2. Methodology

### 2.1. Ethical Consideration

Ethical approval was obtained from the Ethical Review Committee (ERC) of Aga Khan University prior to the initiation of the study with approval number ERC#4794. Informed consent was obtained from all participants or their legal guardians.

### 2.2. Participant Enrollment

The study design and data collection protocols were developed as part of the United World for Antiviral Research Network (UWARN), one of the NIH-funded Centers for Research in Emerging Infectious Disease (CREID). The protocols were designed to create a cohort of patients presenting with clinical symptoms consistent with acute febrile illness (suggestive of Arboviral or other emerging viral infection). These patients were recruited from the inpatient and outpatient settings from The Aga Khan University Hospital from 2021 to 2023 based on the inclusion and exclusion criteria of the study. The majority of subjects belonged to the Karachi region which is one of the most densely populated area of Pakistan (Mean population density 21862). **Figure 1** shows the flow of subject enrollment.

**Figure 1.**
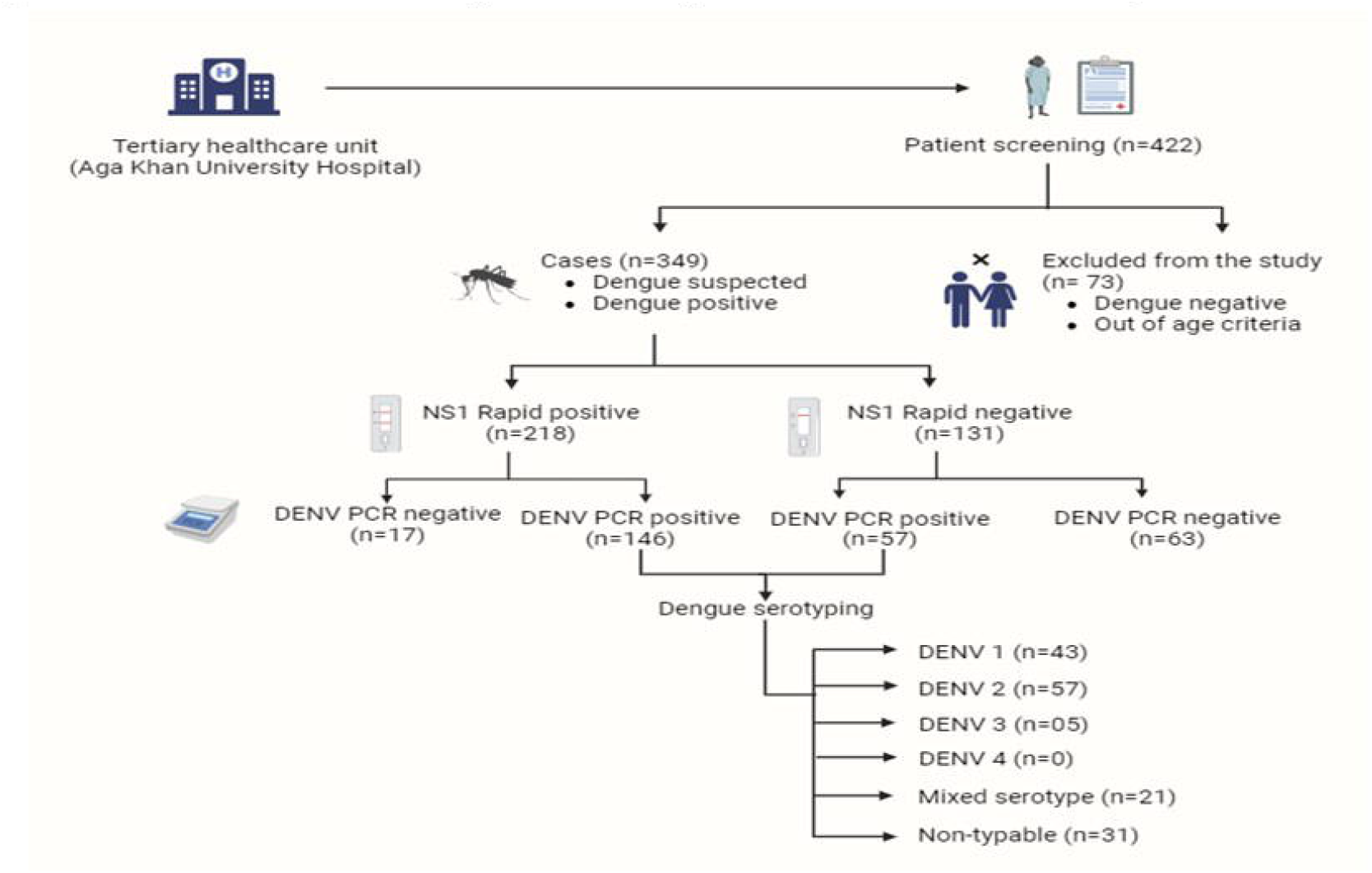
Enrollment and dengue Screening workflow in UWARN study. Enrollment and Dengue Screening workflow in UWARN study. Flow chart of recruitment of dengue cases in UWARN study, Pakistan site. Fever of other origin were not enrolled in our study (n=73)

To define cases for enrollment as confirmed or suspected dengue, we followed clinical symptoms and classification as per World Health Organization (WHO) and Pan American Health Organization (PAHO) guidelines (PAHO World Health Organization, 2017)(14). The criteria include (a) Admission within 1 -2 weeks and clinical suspicion of arbovirus infection or confirmed diagnosis of arboviral disease, (b) Sudden, moderate to high-grade fever (>39° C / 102.2° F), typically of 2 to 7 days duration, (c) any manifestations such as nausea or vomiting, diarrhea, exanthema (non-characteristic, appears between days 5–7), mild to intense pruritus, myalgia, headache, abdominal pain, petechiae/skin bleeding, and dental/gum bleed, (d) patient age less than or equal to 75 years.

Patients not meeting inclusion criteria and those refusing consent were excluded from the study. Age > 75 years and less than 1 year.

### 2.3. Case Definition

The enrolled patients were classified into DENV+ and DENV-based on serological and molecular evidence of DENV.

#### Dengue Positive

Participant with NS1+ or IgM+ or PCR+ (n=287)

#### Dengue Negative

Participant with negative NS1-or IgM- and/or PCR-(n=62)

### 2.4. Clinical Diagnosis

A total of 349 Dengue patients were classified on dengue severity as per WHO criteria into severe dengue (SD; n=5) and non-severe dengue (NSD), which was further divided into dengue with warning signs (DWS; n=39), dengue without warning signs (DWOS; n=261), and with other diagnosis (n=43) One sample did not have record available for classification.

### 2.5. Sample Collection

Blood samples (n=349) were collected in an EDTA tube from each participant enrolled during acute phase of disease. Samples were transported to the laboratory within 30 minutes of collection and tested for NS1 antigen using Abbott Panbio dengue Early Rapid Kit (Suppl. Material) at the time of enrollment followed by sample storage at 4 °C. Plasma was separated and stored at -80 °C.

### 2.6. RNA Extraction

RNA was extracted from the plasma sample using the QIAamp® Viral RNA Extraction Kit (Qiagen-GmbH, Hilden, Germany) following the manufacturer’s protocol. The RNA was eluted in elution buffer and stored at –80°C. The integrity of the extraction reagents and the successful recovery of RNA from clinical samples, the presence of RNase P was confirmed using RNase P probe-based qRT-PCR assay from Biosearch Technologies.

### 2.7. qRT-PCR Amplification

Extracted RNA was subjected to the Pan dengue and dengue serotyping PCR using the Platinum Superscript III Invitrogen One-Step qRT-PCR kit (Invitrogen, Life Technologies, Carlsbad, CA, USA) (15). The Pan dengue and serotyping primers and probes were purchased from Biosearch Technologies (S1A & S1B Table). The positive controls for DENV1, DENV2, DENV3, and DENV4 were acquired from BEI resources, and were used at 1:10 dilution as working concentrations All primers and probes working concentrations were adjusted at 20 µM followed by the addition of RNA template into each reaction. qRT-PCR was performed using CFX96 Bio-Rad (Hercules, CA) platform. The cycling conditions included an initial step at 52°C for 15 minutes followed by 94°C for 2 minutes for the reverse transcriptase reaction and initial denaturation, respectively. The PCR was set up for 45 cycles with denaturation at 94°C for 15 seconds and annealing with acquisition at 55°C for 20 seconds, followed by extension at 60°C for 20 seconds, and a final extension at 68°C for 20 seconds. Fluorophore signals were detected on their respective channels. Any curve crossing the threshold before 42 cycles was considered positive after baseline and threshold adjustments.

### 2.8. Statistical Analysis

Descriptive statistics (Mean ±Standard Deviation) were calculated for quantitative variables. Laboratory values in the clinical setting were assessed in comparison to the normal reference range specific to the corresponding gender as required. For the DENV PCR+ and PCR-samples, Wilcoxon rank-sum test was performed to identify the differences among three groups. Moreover, the Kruskal Wallis test was applied to determine the differences of hematological parameters and Ct value distribution in dengue serotypes. P value < 0.05 was used as a cutoff for statistical significance. All the analysis was performed using R software v.4.2.2.

## 3. Results

### 3.1. Demographics and Clinical Features of Dengue Infection

We screened 422 patients from in-patient and out-patient settings of Aga Khan University and among them, 349 subjects met the inclusion criteria (Fig. 1). Cases with confirmed dengue diagnosis and subjects with dengue-like symptoms (Dengue suspected) were enrolled during the study period (2021 – 2023). Approximately half of the cases (n = 168) were enrolled during Sep-Oct 2022 due to heavy rainfall and floods in Sindh province (Fig. 2A). The enrollment pattern depicts that most cases (n = 85) DENV+ from NS1 were included from Sep-Oct 2022 (Fig. 2B). Initially 218/349 (62.4 %) patients were tested positive for NS1 antigen. The mean age of DENV positive (Participant with NS1+ or IgM+ or PCR+) was 33±14 years and DENV negative (NS1-or IgM- and/ or PCR-) were 31±15 years, including both Male (n = 209) and Female (n =140).

**Figure 2.**
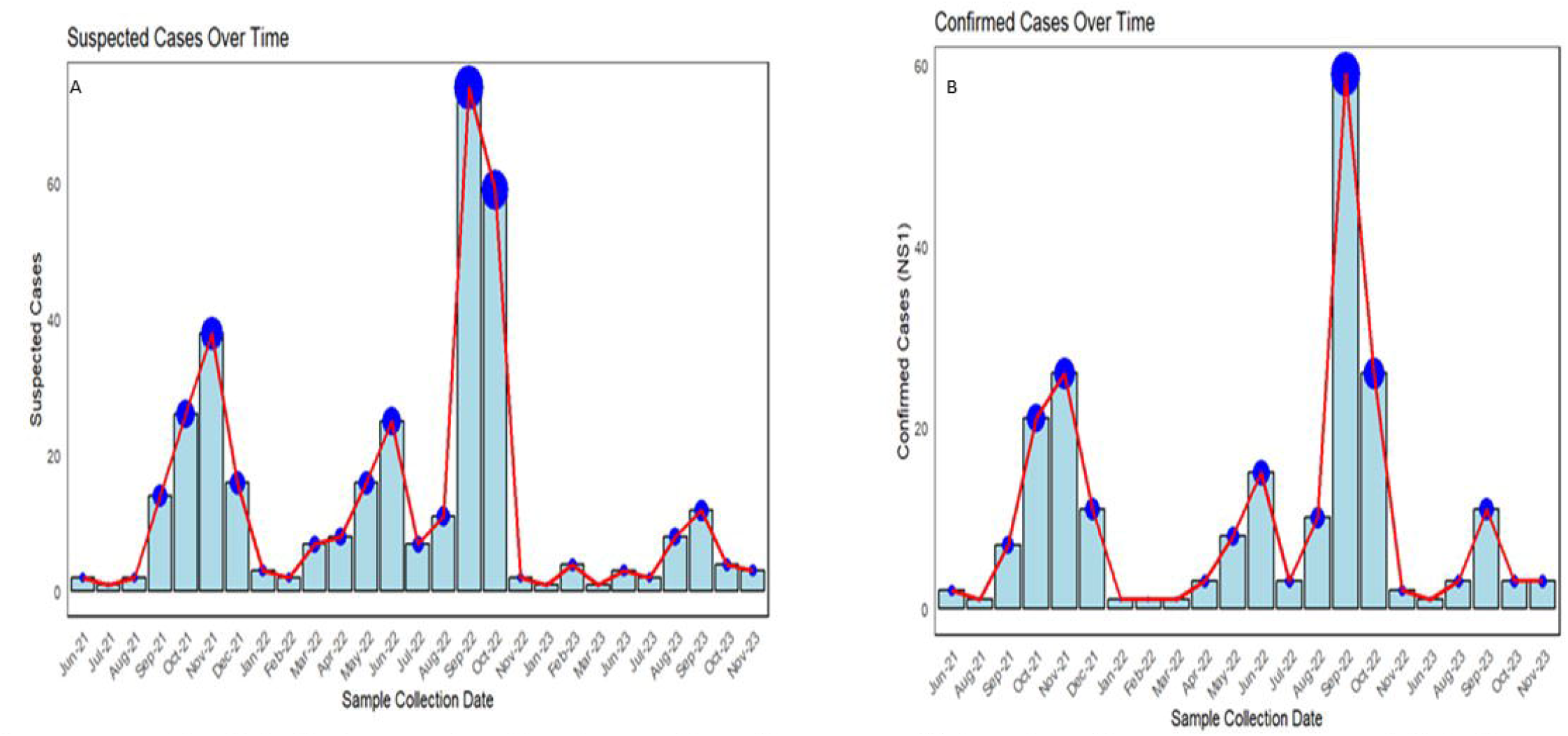
Enrollment of patients in the study over a period of two years. An epidemiological curve showing the number of Suspected (A) and confirmed dengue cases (B) during 2021-2022 according to the diagnosis by NSl antigen; red curve shows the trend of confirmed dengue cases.

Male gender (60%) was found to be predominant in comparison to female gender. Respiration rate (p= 0.005), lymphocyte (p=0.001), neutrophils (P<0.001), platelet count (P<0.001), WBC (p<0.001), ALT (p<0.001), AST (p<0.001) and creatinine (p<0.001) showed statistically significant differences between DENV positive and negative cohorts. In hematological parameters were found to be statistically significant (Table 1).

**Table 1.**
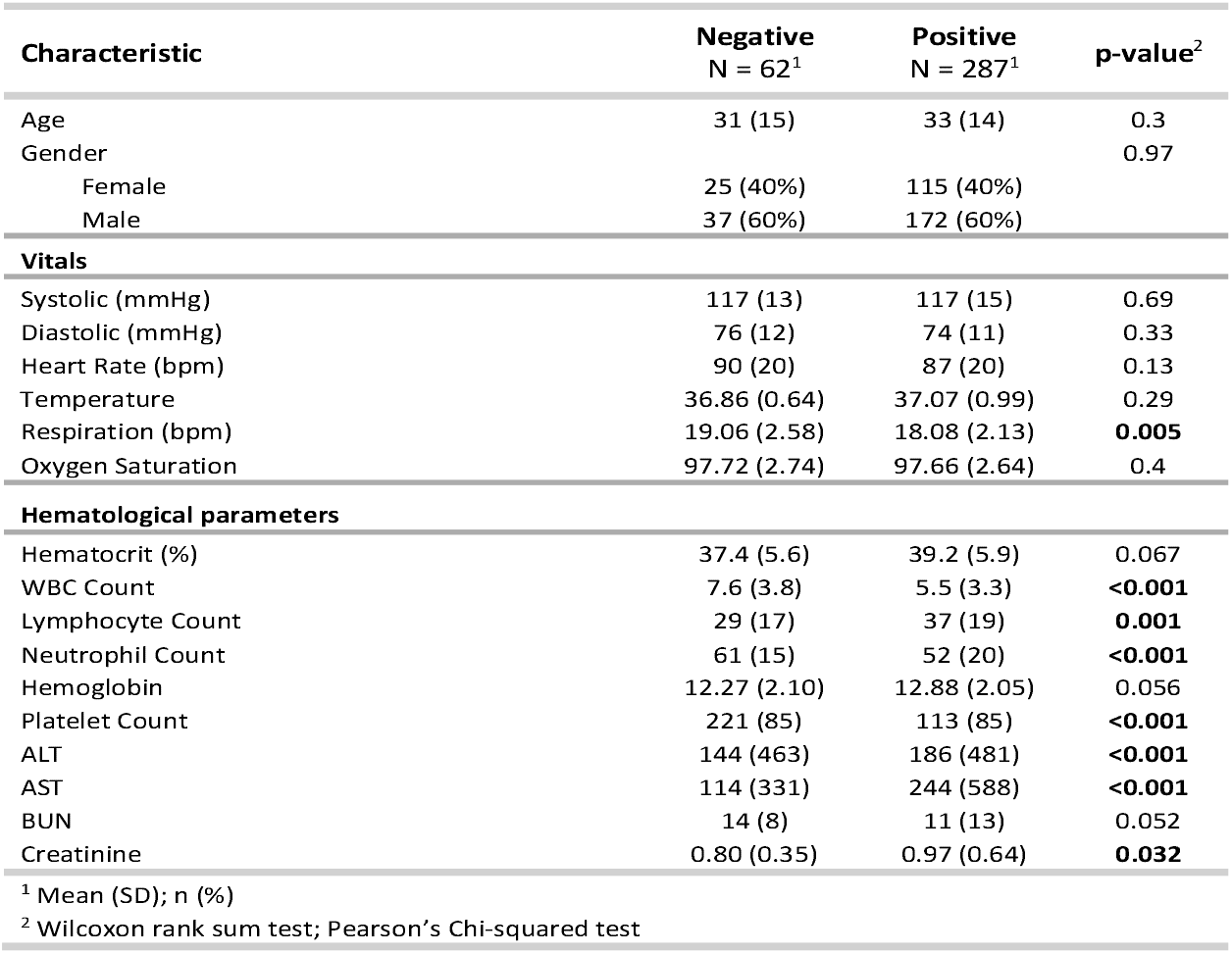
Demographics and clinical characteristics of DENV cohort-clinical definition (N=349)

After the initial screening, Pan-Dengue-PCR was performed on 283/349 subjects. A total of 283 PCR were performed and 66 samples were not processed further for PCR analyses due to limitation of reagents. Of the 283, 203 (71%) were identified as DENV+ through qRT-PCR, and within this subset, 157 individuals were subjected to testing for four DENV serotypes (Fig. S1). The overall screening and algorithm of testing is shown in Fig 1. Among the DENV positive cases as per case definition (PCR+ OR IgM+ OR NS1+) (n=287, male gender was predominant 172 (60%) compared to females 115(40%) (Table 1 & S3 Table).

The clinical signs and symptoms, as classified by the WHO dengue severity were assessed in patients who tested positive for both NS1 and PCR. The history of fever at the time of enrollment was identified as the most common across all three (98%) [dengue without warning signs (DWOS), dengue with warning signs (DWS), and severe dengue (SD)] categories of dengue infection. Moreover, higher percentages of abdominal pain, body aches and diarrhea were observed in severe dengue infection in comparison to other categories (Fig. 3).

**Figure 3.**
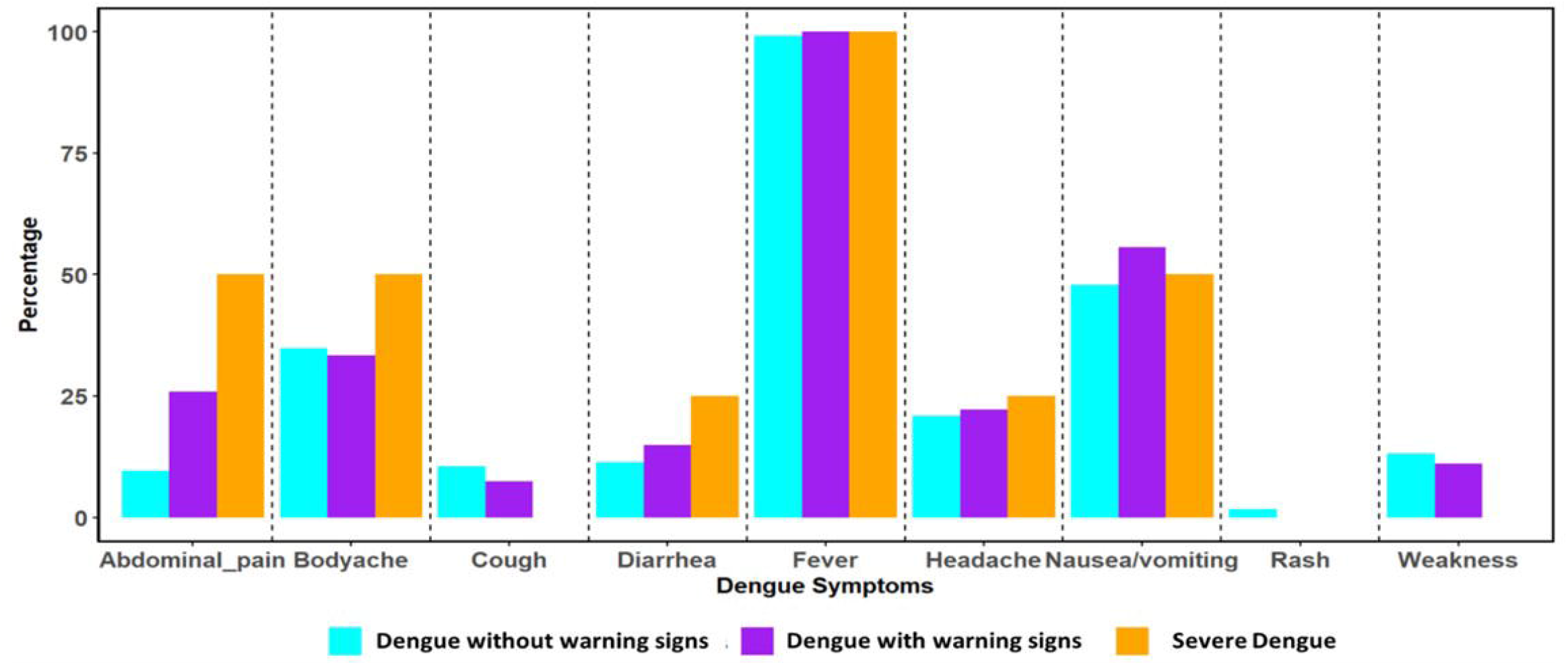
Clinical symptoms in Dengue patients as per WHO criteria. Associated symptoms of dengue infection classified according to WHO criteria. Fever is found to be the most common symptoms among all categories. Orange represents SD; Cyan represents DWWS, and purple represents DWOS.

### 3.2. Sensitivity of Serological Pattern for dengue Infection

The sensitivity of q-RT PCR was calculated based on the based on the clinician diagnosis. Clinical Dx+ includes all dengue fever, dengue-like illness, suspected dengue fever, suspected viral fever, viral meningitis, suspected zika virus. Clinical Dx-includes other than dengue/viral fever and with other comorbidities as mentioned in suppl. Appendix I sheet B & S3 table. This constitutes a total of 349 cases which include Dx+ (n=287) [NS1+/IgM+/PCR+] and Dx – [NS1-/IgM-/PCR-] (n=62) groups. The sensitivity of clinical diagnosis compared to PCR was found to be 87.25%, accompanied by a specificity of 68.35%, a Positive Predictive Value (PPV) of 98.13%, and a Negative Predictive Value (NPV) of 22.01%, as detailed in Table 2 & S4 Table.

**Table 2.** Sensitivity of Clinical Diagnosis of DENV with Gold Standard diagnostics.

| Statistics | Value (%) | 95% Confidence Interval (CI) |
| --- | --- | --- |
| Sensitivity | 87.25 | 81.89 – 91.50 |
| Specificity | 68.35 | 56.92 – 78.37 |
| Positive Predictive Value (PPV) | 98.13 | 97.42 – 98.64 |
| Negative Predictive Value (NPV) | 22.01 | 16.06 – 29.41 |

Besides, 41 cases presented with multiple unrelated symptoms secondary to viral illness. The clinical presentations were shown in suppl. Appendix I Sheet B. These samples were also subjected to q-RT PCR where initial NS1 or IgM tests were negative on screening (n=41). We found 19 samples to be positive for dengue PCR and its serotypes (DENV 1= 10, DENV 2=7, DENV 1/DENV 2=2). This was an additional finding in our study which was picked by highly sensitive PCR test. A total of 46% positivity was observed for RT PCR results.

### 3.3. Serotype-specific Detection of dengue Virus

Of 203 qRT-PCR positive cases, 157 were subjected to DENV serotyping. The mean Ct values for DENV 1 was 31.05, DENV 2 was 32.95, DENV 3 was 33.81 and mixed serotype was 32.16. Moreover, no significant differences were observed in the mean Ct values between all the groups (DENV 1, DENV 2, DENV 3 and mixed serotypes) (S2 Fig & S2 Table). Among 157, DENV 1 (n = 43) was 27% and DENV 2 (n=57) was 36%, such that DENV 1 and DENV 2 were the predominant serotypes. Moreover, DENV 3 accounted for only 3% (n = 5), and DENV 4 was not detected through serotype specific PCR analysis. Additionally, 13% of cases (n = 21) were identified as mixed serotypes (Fig. S1 Panel B, Suppl. Appendix Sheet C). Moreover, there were also a few non-typeable samples 20% (n =31), which were not classified using primer set.

### 3.4. Clinical Presentation in dengue Infection

Clinical parameters, including vitals and hematological parameters, were available for the 349 enrolled patients at the time of enrollment. The mean vitals values were similar between dengue PCR positive and negative patients except for temperature parameter (Table 3). Similarly, the majority of DENV patients exhibited mean normal body temperature, however a proportion of patients (31% DENV+ and 16% DENV-) showed slightly raised body temperature (>37.0°C). The observed differences were statistically significant (p = 0.003). Abnormal respirations (>20 breaths per minute) were noted in 9% of individuals with DENV+ and 6% with DENV-. Moreover, oxygen saturation was normal in all cases (i.e., 100% for DENV+ and 98.5% for DENV-). The details of the normal and deranged vitals for both groups are mentioned in Suppl. Appendix I Sheet D.

**Table 3.** Demographics and clinical characteristics of qRT-PCR confirmed cases.

| Characteristic | Negative<br>N = 80<sup>1</sup> | Positive<br>N = 203<sup>1</sup> | p-value<sup>2</sup> |
| --- | --- | --- | --- |
| Age | 31 (1 - 70) | 34 (3 - 75) | 0.28 |
| Gender |  |  | 0.37 |
| Female | 30 (38%) | 88 (43%) |  |
| Male | 50 (63%) | 115 (57%) |  |
| **Vitals** |  |  |  |
| Systolic (mmHg) | 118 (84 - 152) | 117 (84 - 167) | 0.45 |
| Diastolic (mmHg) | 75 (50 - 112) | 74 (45 - 109) | 0.37 |
| Heart Rate (bpm) | 89 (13 - 182) | 85 (10 - 145) | 0.3 |
| Temperature | 36.80 (36.00 - 39.50) | 37.12 (34.00 - 41.20) | **0.013** |
| Respiration (bpm) | 18.46 (15.00 - 29.00) | 18.07 (12.00 - 29.00) | 0.11 |
| Oxygen Saturation | 97.45 (84.00 - 100.00) | 97.74 (90.00 - 100.00) | 0.58 |
| **Hematological Parameters** |  |  |  |
| Hematocrit (%) | 38.4 (22.8 - 51.4) | 38.3 (23.3 - 53.4) | 0.76 |
| WBC Count (10<sup>9</sup>/L) | 7.1 (1.3 - 20.0) | 5.6 (1.0 - 21.9) | **<0.001** |
| Lymphocyte (%) | 36 (4 - 97) | 37 (5 - 97) | 0.43 |
| Neutrophils (%) | 54 (16 - 91) | 52 (9 - 91) | 0.34 |
| Hemoglobin (g/dL) | 12.52 (7.30 - 16.60) | 12.69 (7.80 - 17.50) | 0.64 |
| Platelet Count (10<sup>9</sup>/L) | 186 (12 - 515) | 109 (6 - 479) | **<0.001** |
| ALT (IU/L) | 280 (10 - 5,241) | 163 (9 - 3,515) | 0.34 |
| AST (IU/L) | 165 (21 - 1,722) | 260 (8 - 6,414) | **0.039** |
| BUN (mg/dL) | 12 (3 - 39) | 12 (3 - 139) | 0.16 |
| Creatinine (mg/dL) | 0.86 (0.20 - 2.20) | 0.98 (0.30 - 6.30) | 0.28 |

<sup>1</sup> Mean (Min - Max); n (%)
<sup>2</sup> Wilcoxon rank sum test; Pearson's Chi-squared test

The hematological parameters of DENV+ and DENV-(n=283) patients showed low platelet count, elevated AST, and deranged ALT levels (Table 3). The mean platelet counts within 1-2 days of enrollment for DENV+ patients were below the normal range of 150 – 400 × 10^9/L. Similarly, ALT and AST levels were also found to be deranged from the normal range (≤40). The percentages of the hematological parameters and comparison with normal ranges is shown in in Suppl. Appendix I Sheet D. Furthermore, low platelets (p<0.0001) and WBC count (p = 0.001) were observed in DENV+ group (Table 3 and Appendix Sheet D). In addition, the differences in hematological profiles of DENV serotype specific groups (DENV 1, DENV 2, DENV 3 and mixed) were also measured showing statistically significant differences for Creatinine (p=0.022) (S5a Table). The difference in hematological parameters were observed for DENV 1 and DENV 2 serotypes, with Platelets count being lower (123 vs. 93 10^9/L; p=0.025), while higher Creatinine level in DENV 2 serotype (0.76 vs 1.10 mg/dL; p=0.002) (S5b Table). Additionally, the hematological parameters in serotype positive group (n=126) were compared as per WHO criteria of DWWS and DWOS. In this analysis WBC (p=0.024), Platelets (p=0.006) and Creatinine (p=0.007) were found to be significant (S6 Table). As per WHO criteria, 261 were classified as DWOS, 39 with DWWS and 5 as SD. This makes a total of 305 patients, 43 did not classify in any of the above categories, but presented with other clinical presentations. The median levels of AST (p=0.016) and ALT (p=0.009) were significantly higher in patients with SD compared to those with NSD (Table 4). A proportion of 49.8% (152/305) patients had abnormal ALT levels (>40 U/L), and 51.4% (157/305) of patients had abnormal AST levels (>40 U/L). ALT levels >1000 U/L were observed in six patients, two had dengue shock syndrome and one of them had co-infected with hepatitis A virus. Renal dysfunction was also significantly higher in severe dengue cases in comparison to NSD with elevated BUN (p=0.005) and creatinine (p <0.001) (Table 4). The SD group (n=5) presented with Dengue shock syndrome, Acute Kidney injury, Multiorgan dysfunction (MOD), Septic shock, and Dengue Hemorrhagic fever.

**Table 4.** Clinical parameters and their significance among dengue severity.

| Characteristic | Dengue with warning signs<br>N = 39<sup>1</sup> | Dengue without Warning Signs<br>N = 261<sup>1</sup> | Severe DENGUE<br>N = 5<sup>1</sup> | p-value<sup>2</sup> |
| --- | --- | --- | --- | --- |
| Hematocrit | 37.8 (5.7) | 39.3 (5.9) | 33.6 (8.5) | 0.11 |
| WBC Count | 5.7 (2.5) | 5.5 (3.6) | 7.9 (4.1) | 0.12 |
| Lymphocyte Count | 40 (17) | 37 (19) | 18 (5) | **0.049** |
| Neutrophil Count | 47 (19) | 53 (20) | 75 (8) | **0.014** |
| Hemoglobin | 12.44 (1.86) | 12.91 (2.09) | 11.13 (1.94) | 0.086 |
| Platelet Count | 82 (75) | 125 (89) | 179 (103) | **0.001** |
| ALT | 239 (442) | 132 (244) | 1,929 (2,324) | **0.009** |
| AST | 428 (1,119) | 173 (234) | 1,599 (2,022) | **0.016** |
| BUN | 11 (10) | 11 (13) | 26 (10) | **0.005** |
| Creatinine | 0.92 (0.65) | 0.89 (0.28) | 3.58 (2.21) | **<0.001** |
| Days of Hospitalization | 2.64 (2.22) | 1.65 (2.86) | 5.40 (5.55) | **0.002** |

<sup>1</sup> Mean (SD)
<sup>2</sup> Kruskal-Wallis rank sum test

Thrombocytopenia was further categorized into grade 0 (>150,000), grade I (75,000 – 150,000), grade II (50,000 – 75,000), grade III (25,000-50,000), and grade IV (< 25,000). Notably, there were significant differences in thrombocytopenia with respect to the days of illness (p = 0.03) and hospitalization (p = 0.001) (Fig. 4 panel A and B). Increased Days of illness and hospitalization was also observed in patients with higher grade of thrombocytopenia. In addition, the dengue severity groups also showed statistically significant differences with the hospitalization (p = 0.001) as shown in Fig. 4C. Additionally, the neutrophil-to-lymphocyte ratio (NLR) was also found to be significantly deranged (NLR > 3.0) in SD patients.

**Figure 4.**
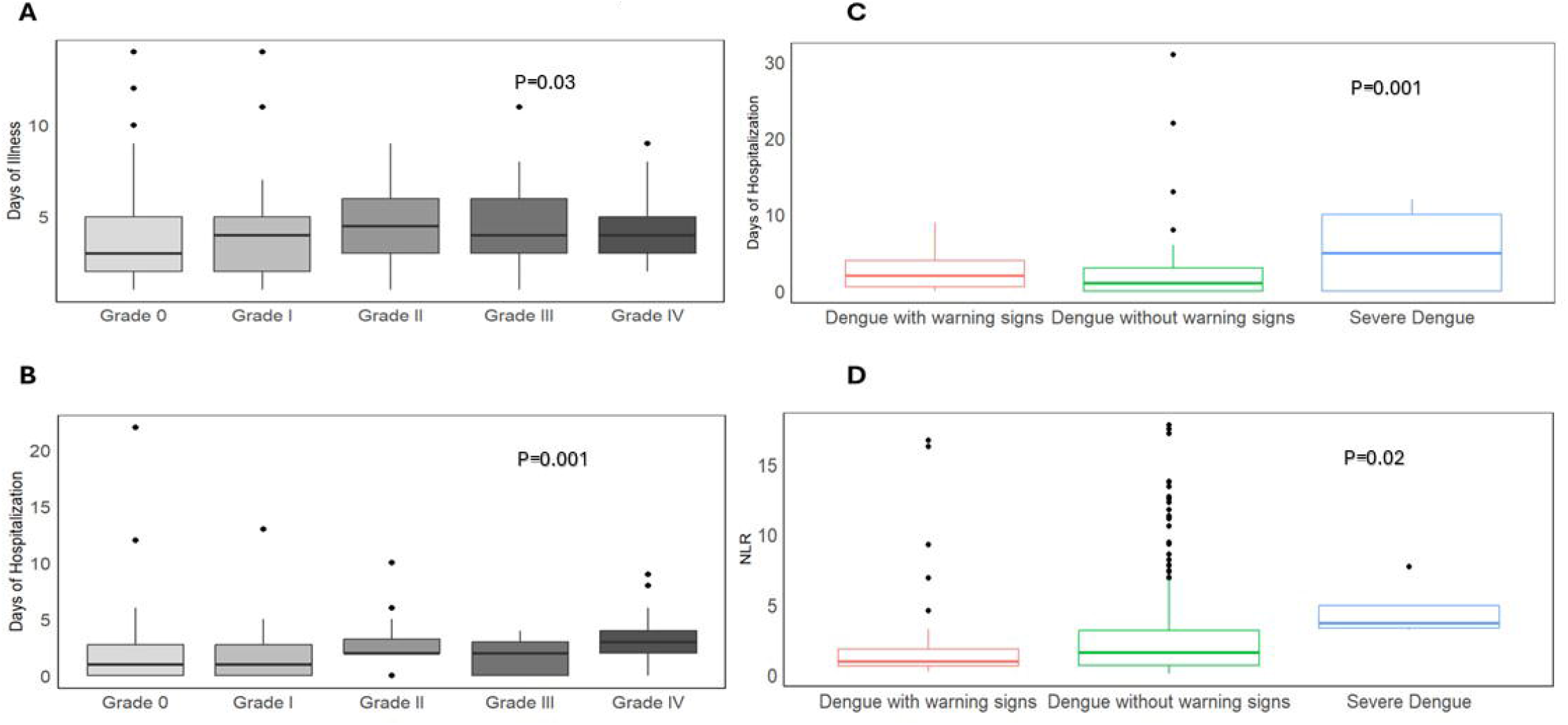
Comparison of platelet grading and dengue severity groups with days of illness, hospitalization, and Neutrophil-to-Lymphocyte Ratio (NLR). Comparison of platelet grading and dengue severity groups with days of illness, hospitalization, and Neutrophil-to-Lymphocyte Ratio (NLR).(A) Association between days of illness and grade of platelet grading.(B) Association between hospitalization and platelet grading. (C) Association between dengue severity groups and hospitalization. (D) Association between dengue severity groups and NLR

### 3.5. Clinical Diagnosis of Dengue Mixed Serotypes

Of those PCR positive (n=203), only 157 were subjected to serotyping assay, 46 samples were not processed further due to limitation of reagent. 126/157 were positive with any one serotype of Dengue, and 31 was non typeable. Of these 126 cases 30 (23.8%) were initially negative for NS1 but later found to be DENV PCR+. Among those, 21 had Dengue fever/ Dengue like illness/ viral fever without any other complication, while 9 had Dengue fever with some complications involving GI symptoms (n=1), upper respiratory tract symptoms (n=2), and other complications such as metabolic acidosis (n=1), drug reaction(n=1), pancytopenia(n=1), UTI (n=1), suspected Zika virus infection (n=1) and connective tissue disorder(n=1). Of those 31 Non typeable serotypes, 13 had a diagnosis of dengue fever, 9 had a diagnosis of fever and viral fever, 7 were diagnosed with enteric fever while 2 were diagnosed with tonsilitis and community acquired pneumonia. These diagnoses were probably not correct, as they were positive for Pan Dengue PCR (Suppl. Appendix I Sheet B). In addition, mixed serotype infection included (n=21) cases, in which DENV 1|DENV 2 is predominant (n=20) and only 01 case was found with DENV 1| DENV 3 co-infection. Of these, 13/21 were in-patients, while the remaining 8 were out-patients.

Among the mixed serotype cases, 20 individuals were diagnosed with dengue fever/fever/suspected dengue fever/viral fever and 1 case was diagnosed with upper respiratory tract infection (Suppl. Appendix I Sheet A). Moreover, within mixed serotypes, a significant majority of cases were categorized as dengue without warning signs (18/21) (S7 Table). Further to this, it has been observed that the majority of patients in DENV 2 and mixed serotypes were hospitalized patients (74%). Only 05 patients were identified as DENV 3 and all of them were in-patient as shown in S3 Fig.

## 4. Discussion

The first Dengue outbreak in Pakistan was reported in 1994, followed by significant outbreaks in 2005, 2011, 2017, 2018, 2019, and 2022(8). The recent outbreak reported 75,450 Dengue cases between January and November 2022 (Data from Pakistan’s National Institute of Health) as huge economic and health crises exacerbated by flooding. Pakistan ranks 30^th^ in the climate change performing index (CCPI) and has received a low score in the Climate Policy category, indicating vulnerabilities to climate change impacts, including increased dengue incidence. (https://ccpi.org/country/pak). Our study focused on the epidemiological characteristics of dengue virus infection among hospitalized and out-patient cohorts in Karachi, emphasizing the monsoon season of 2022. Most cases were enrolled between September – October 2022, a period marked by heavy rainfall and catastrophic flooding(9). Moreover, August 2022 stands out as the wettest month in the past 50 years, and October 2022 had the highest number of dengue cases reported in Pakistan. The correlation between high precipitation levels and dengue incidence is evident, with flood-affected regions like Sindh experiencing a higher prevalence of infections (6, 17). The urban areas such as Karachi, with dense populations and inadequate sewage and drainage systems faced significant challenges in controlling mosquito breeding sites, further contributing to the spread of dengue(2, 18).

Demographic analysis revealed a predominance of male cases, due to higher outdoor exposure or differences in health seeking behavior, influenced by social and cultural factors(19). Adults aged 16-45 years were most frequently affected, consistent with previous studies which also reported this age group to be more vulnerable to viral illness which might be due to their increased exposure to outdoor activities(13).

Serotype analysis identified DENV 1 and DENV 2 as primary serotypes which are consistent with previous reports from Pakistan (19) and neighboring countries. Notably, the 2006 outbreaks in Karachi and New Delhi were dominated by DENV 2 and DENV 3, which resulted in numerous fatalities(20). Data from 2017 to 2022 indicate fluctuating dengue incidence, with significant spikes in 2019 and 2021 and a substantial rise in 2022, particularly in Khyber Pakhtunkhwa, Sindh, and Punjab.

Comparative studies in other regions such as Jaipur, Rajasthan and Surabaya, Indonesia, showed varying serotypes distribution and clinical outcomes. In a study conducted in a tertiary care hospital in Jaipur, Rajasthan, the most prevalent dengue serotypes were DENV 2 and DENV 3 while severe cases were associated with DENV 2 and DENV 4 infection (21). A study conducted in Surabaya, Indonesia, correlating serotypes with clinical and laboratory data, found no significant differences except for a higher lymphocyte count in DENV 1 infected patients (22).

A retrospective study in Malaysia noted more severe clinical manifestations in co-infected patients(10). Regarding clinical features, most DENV cases in our study presented with fever, nausea or vomiting, body ache and headache, consistent with findings across the country (17). Retro orbital pain and maculopapular rash were less common. No significant vital differences were observed between PCR-positive and negative patients, except for higher temperatures in positive cases due to viral inflammation.

The hematological parameters of patients with and without DENV infection are seen within normal ranges, with a few exceptions for platelet count, ALT, and AST. DENV positive patients showed decreased platelet counts, aligning with other studies indicating significant declines during severe dengue cases. Elevated ALT and AST levels in both DENV positive and negative cases were consistent with previous reports from Rawalpindi, Pakistan, indicating liver involvement during dengue infection(23). Liver involvement is characterized by hepatomegaly and abnormal liver enzymes, and is frequently observed during dengue infection, particularly in patients with dengue hemorrhagic fever (DHF). Moreover, hepatomegaly, coupled with elevated liver enzymes, was notably more prevalent in DHF patients. Several studies have proposed the use of liver enzymes (ALT and AST) as indicators to assess disease severity in dengue patients, as markedly elevated liver enzymes serve as an early warning sign for severe disease and clinical bleeding (24). Higher grade of thrombocytopenia in DENV also suggested marker for duration of hospital stay and days of illness (25).

In summary, the current study conducted in a large tertiary care center in Pakistan, highlights the profound impact of Dengue worsened by flooding and poor sanitation, posing considerable challenges to public health. The high prevalence of Dengue cases in flood-prone regions emphasizes the urgent need to address the health impacts of climate change. Furthermore, the rising Dengue rates in urban centers like Karachi underscore the importance of robust infrastructure and sanitation. Immediate measures by authorities are crucial to control vector populations and protect public health. The main limitation of this study includes lower number of dengue severe cases and the unavailability of the resources to perform dengue sequencing for mixed infections.

## Data Availability

All data presented available in supplementary appendix

## Acknowledgement

Field staff, data management unit staff, patients and families for their contribution, BEI resource for provision of positive controls for PCR, Scott Weaver UTMB WRCEVA for the provision of positive controls, CREID network at NIH for funding and technical support.

