## Supplementary figures and images for "Clinical characteristics and serotype association of dengue and dengue like illness in Pakistan"

### Suppl Fig 2

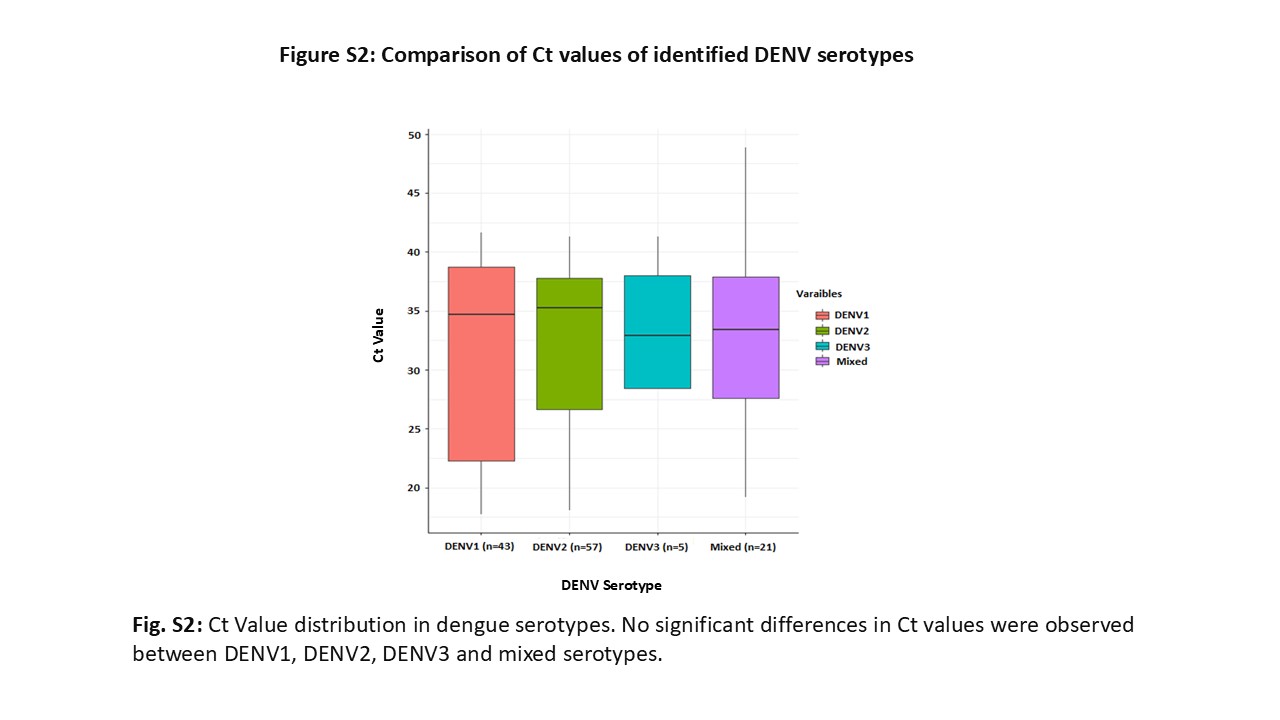

### Suppl. Fig 3

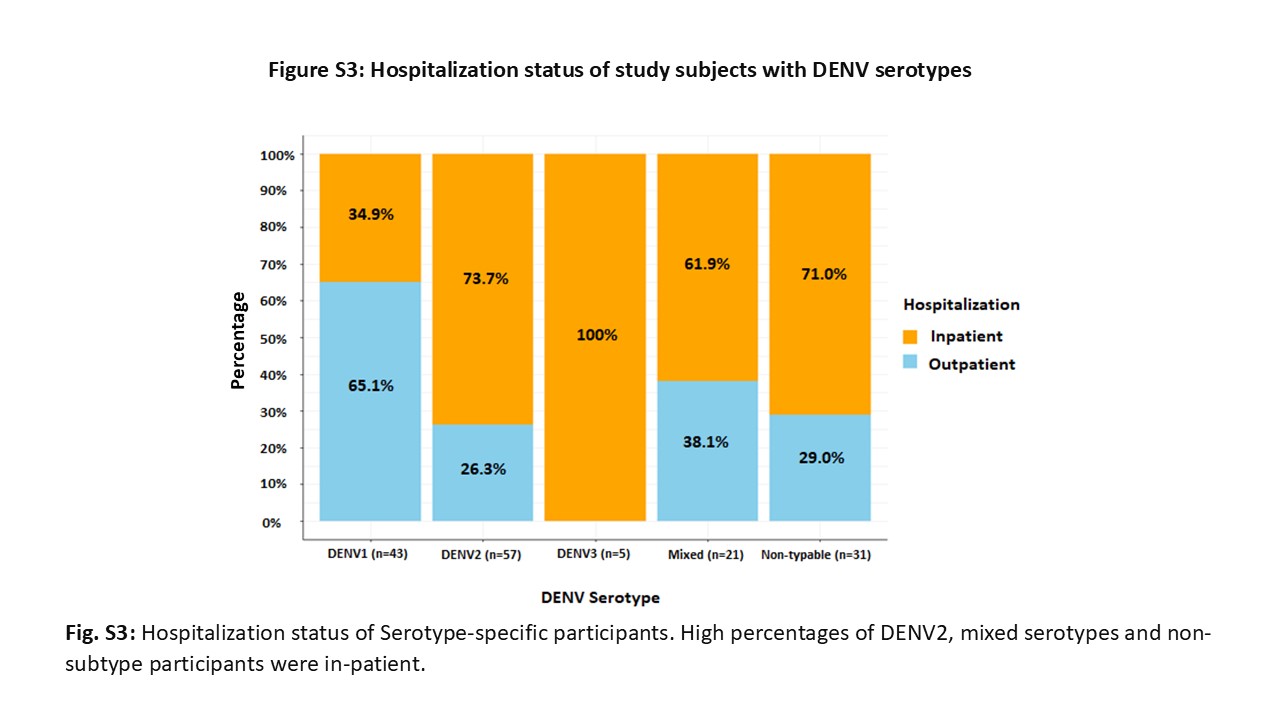

### Suppl.Fig 1

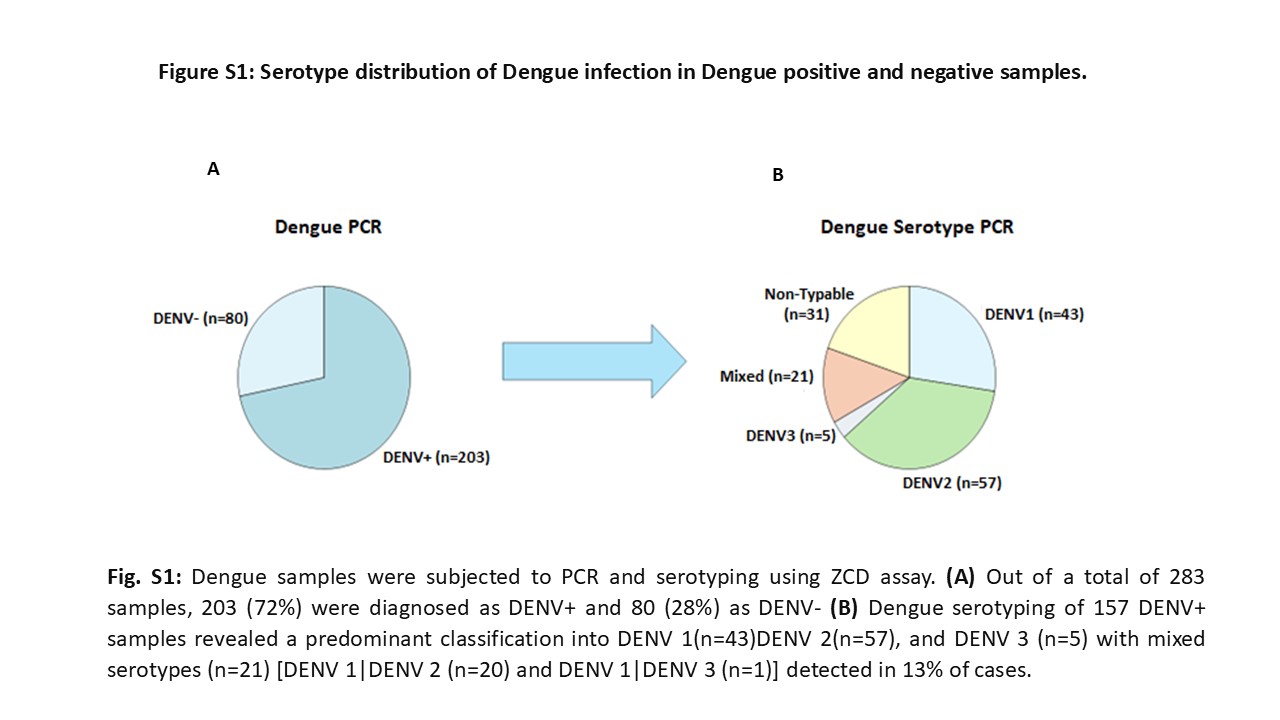
