## Supplemental tables 1-7 for "Clinical characteristics and serotype association of dengue and dengue like illness in Pakistan"

**Supplementary Material Tables**

**Table S1A:** ZCD assay primers for Dengue detection

| Primers | Sequence (5’-3’ d) | Genomic Location | Total Bases | MW |
| --- | --- | --- | --- | --- |
| DV1,2,3-F | CAGATCTCTGATGAACAACCAACG | 86-109 | 24 | 7291.60 |
| DV2-F | CAGATCTCTGATGAATAACCAACG | 87-110 | 24 | 7306.62 |
| DV3-F | CAGATTTCTGATGAACAACCAACG | 85-108 | 24 | 7306.62 |
| DV4-F | GATCTCTGGAAAAATGAAC | 81-99 | 19 | 5826.72 |
| DV1,3-R | TTTGAGAATCTCTTCGCCAAC | DENV-1: 199-179 DENV-3: 198-178 | 21 | 6336.00 |
| DV2-R1 | AGTTGACACGCGGTTTCTCT | 171-152 | 20 | 6079.83 |
| DV2-R2 | AGTCGACACGCGGTTTCTCT | 171-152 | 20 | 6064.81 |
| DV4-R | AGAATCTCTTCACCAACC | 190-173 | 18 | 5370.40 |

**Table S1B:** ZCD assay probes for Dengue detection

| Probes | Sequence (5’-3’) | 5’Fluor | 3’Quencher | Total Bases | MW |
| --- | --- | --- | --- | --- | --- |
| DV1-P | pdCpdUpdCGpdCGpdCGpdUpdUpdUpdCAGpdCApdUApdUA | FAM | BHQ-1 *plus* | 7 | 7198.89 |
| DV2-P | pdCpdUpdCpdUpdCGpdCGpdUpdUpdCAGpdCApdUApdU | FAM | BHQ-1 *plus* | 6 | 7197.89 |
| DV3-P | pdCpdUpdCpdUpdCApdCGpdUpdUpdCAGpdCApdUApdUpdUG | FAM | BHQ-1 *plus* | 7 | 7837.29 |
| DV4-P | pdCpdUpdCApdCGpdCGpdUpdUpdCAGpdCApdUApdU | FAM | BHQ *plus* | 7 | 7182.89 |

**Table S2:** Dengue serotyping specific probes in real time qRT-PCR

| Probes | Sequence (5’-3’) | | 5’Fluor | 3’Quencher | Total Bases | MW |
| --- | --- | --- | --- | --- | --- | --- |
| DV1-P | | CGCGATCTTCAGCATATTGGAAAGACGGTCGGATCGCG | FAM | BHQ-1 | 37 | 12468.02 |
| DV2-P | | CGCGATCGCGTTTCAGCATATTGAAAGGCGGATCGCG | CAL Flour Orange 560 | BHQ-1 | 37 | 12507.17 |
| DV3-P | | CGCGATCCACGCGTTTCAGCATATTGATAGGATCGCG | CAL Flour Red 610 | BHQ-2 | 37 | 12520.19 |
| DV4-P | | CGCGATCTTTCAGCATATTGAAAGGTGGTCGATCGCG | Quasar 670 | BHQ-2 | 37 | 12585.32 |

Table S3: Lab Diagnosis of samples using three diagnostic tests

| **S #** | **NS1** | **IgM** | **PCR** | **Total (n)** | **Diagnosis (Dx)** | **Comments** |
| --- | --- | --- | --- | --- | --- | --- |
| 1 | +ve | +ve | +ve | 5 | Dx +ve |  |
| 2 | +ve | +ve | -ve | 3 | Dx +ve |  |
| 3 | +ve | -ve | +ve | 6 | Dx +ve |  |
| 4 | +ve | -ve | -ve | 0 | **­-** |  |
| 5 | -ve | +ve | +ve | 16 | Dx +ve |  |
| 6 | -ve | -ve | +ve | 17 | Dx +ve |  |
| 7 | -ve | -ve | -ve | 18 | Dx -ve |  |
| 8 | -ve | +ve | -ve | 9 | Dx +ve |  |
| 9 | +ve | N/A | -ve | 14 | Dx +ve |  |
| 10 | +ve | N/A | +ve | 135 | Dx +ve |  |
| 11 | -ve | N/A | +ve | 24 | Dx +ve |  |
| 12 | N/A | +ve | +ve | 0 | **­-** |  |
| 13 | N/A | -ve | +ve | 0 | **­-** |  |
| 14 | N/A | +ve | -ve | 0 | **­-** |  |
| 15 | N/A | N/A | +ve | 0 | **­-** |  |
| 16 | N/A | N/A | -ve | 1 | Dx -ve | Excluded |
| 17 | N/A | -ve | N/A | 1 | Dx -ve | Excluded |
| 18 | N/A | +ve | N/A | 0 | **­-** |  |
| 19 | +ve | +ve | N/A | 0 | **­-** |  |
| 20 | -ve | -ve | N/A | 2 | Dx -ve |  |
| 21 | -ve | +ve | N/A | 3 | Dx +ve |  |
| 22 | N/A | N/A | N/A | 0 | **­-** |  |
| 23 | +ve | N/A | N/A | 51 | Dx +ve |  |
| 24 | -ve | N/A | -ve | 36 | Dx -ve |  |
| 25 | -ve | N/A | N/A | 6 | Dx -ve |  |
| 26 | +ve | -ve | N/A | 4 | Dx +ve |  |
| Total | | | | 351 |  |  |

**Table S4:** Counts of Dengue qRT-PCR with reference to Clinical Diagnosis

| **Dengue Diagnosis*** | **Clinical Dx+** | **Clinical Dx-** | **Total** |
| --- | --- | --- | --- |
| **PCR+** | 178 | 25 | 203 |
| **PCR-** | 26 | 54 | 80 |
| **Total** | 204 | 79 | 283 |

*Clinical Dx+ includes all dengue fever, dengue-like illness, suspected dengue fever, suspected viral fever, viral meningitis, Zika virus. Clinical Dx- includes other than dengue/viral fever and with other comorbidities as mentioned in suppl. Appendix I sheet B.

**Table S5a:** Significance of laboratory parameters among different serotypes.

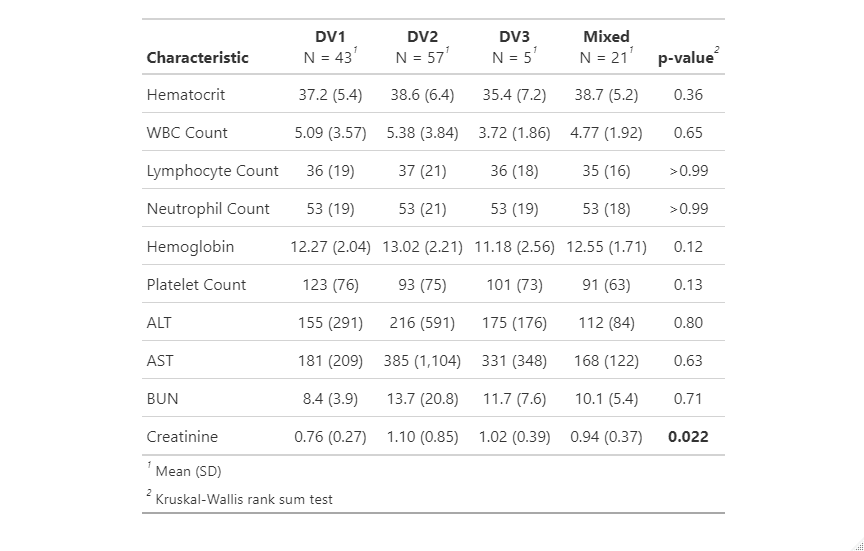

**Table S5b:** Significance of laboratory parameters among DV1 & DV2 serotypes.
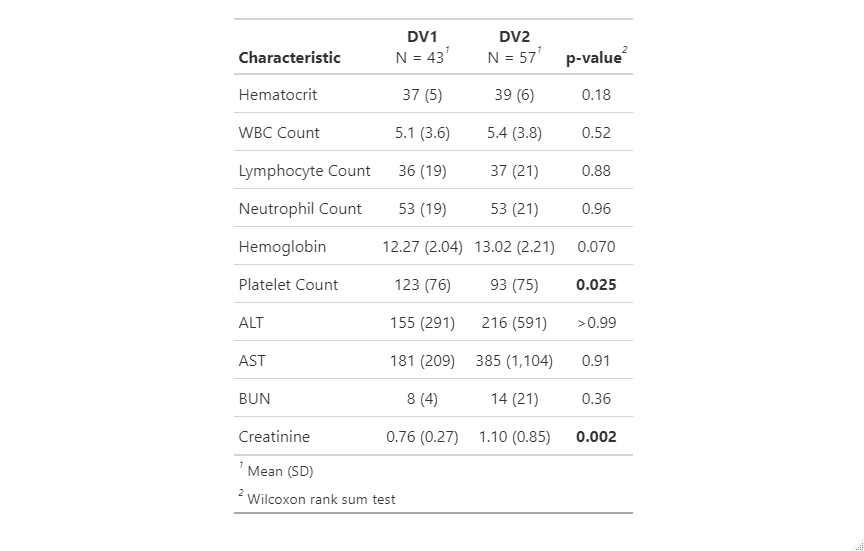

**Table S6: Dengue disease severity in serotype positive samples (n=126)**

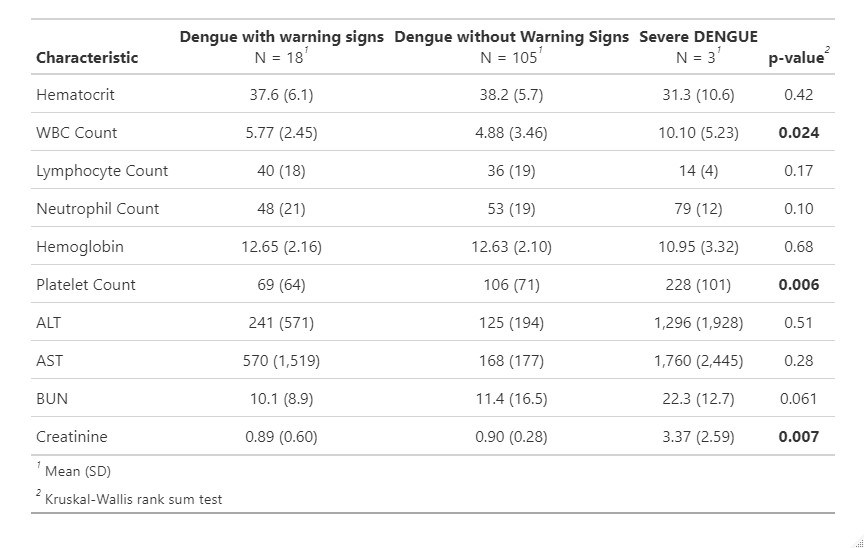

**Table S7:** Clinical diagnosis of dengue serotypes (n=126)

| **Serotypes (n=126)** | **Severe Dengue** | **Dengue with warning signs** | **Dengue without warning signs** |
| --- | --- | --- | --- |
| DENV-1 (n=43) |  | 04 | 39 |
| DENV-2 (n=57) | 03 | 10 | 44 |
| DENV-3 (n=5) |  | 01 | 04 |
| DV1\|DV2 (n = 20) |  | 03 | 17 |
| DV1\|DV3 (n = 1) |  |  | 1 |
